# Prehospital triage of intracranial hemorrhage and anterior large vessel occlusion ischemic stroke: the value of the rapid arterial occlusion evalution

**DOI:** 10.1101/2023.02.24.23286437

**Authors:** Luuk Dekker, Victor J. Geraedts, Jeroen Hubert, Dion Duijndam, Marcel D.J. Durieux, Loes Janssens, Wouter A. Moojen, Erik W. van Zwet, M.J.H. Wermer, Nyika D. Kruyt, Ido R. van den Wijngaard

## Abstract

**Background:** The Rapid Arterial oCclusion Evaluation (RACE) score can identify patients with anterior circulation large vessel occlusion (aLVO) ischemic stroke for transportation to a comprehensive stroke center for endovascular thrombectomy. However, patients with intracranial hemorrhage (ICH) may also benefit from direct transportation to a comprehensive stroke center for neurosurgical treatment. We aimed to assess if the RACE score can distinguish patients with ICH in addition to aLVO stroke from other suspected stroke patients.

**Methods:** We analyzed data from the Leiden Prehospital Stroke Study: a multicenter, prospective, observational cohort study in two Dutch ambulance regions. Ambulance paramedics documented prehospital observations in all patients ≥18 years with suspected stroke. We calculated the sensitivity, specificity, positive predictive value (PPV) and negative predictive value (NPV) of a positive RACE score (≥5 points) for a diagnosis of ICH or aLVO stroke, compared to patients with non-aLVO stroke, TIA, or stroke mimic. Additionally, we performed a multivariable logistic regression analysis and calculated adjusted odds ratios (aOR).

**Results:** We included 2004 stroke code patients, of whom 149 had an ICH, 153 an aLVO stroke, 687 a non-aLVO stroke, 262 a TIA and 753 a stroke mimic. Patients with ICH and aLVO stroke more often had a positive RACE score than other suspected stroke patients (respectively 46.2% and 58.0% vs. 6.4%, p<0.01). A positive RACE score had a sensitivity of 52.7%, specificity of 93.6%, PPV of 55.4% and NPV of 92.9% for a diagnosis of ICH or aLVO stroke. In multivariable analysis, a positive RACE score had the strongest association with ICH or aLVO stroke (aOR 10.11, 95% CI 6.84-14.93).

**Conclusions and Relevance:** Our study shows that the RACE score can also identify patients with ICH in addition to aLVO stroke. This emphasizes the potential of the RACE for improving prehospital triage and allocation of stroke patients.

## INTRODUCTION

Stroke is a leading cause of death and disability worldwide.^1^ The majority of strokes are ischemic, which can be treated with intravenous thrombolysis (IVT) and endovascular thrombectomy (EVT). The efficacy of both IVT and EVT is highly time sensitive.^2-4^ IVT can be administered in all stroke centers, but is less effective in patients with an underlying anterior circulation large vessel occlusion (aLVO).^5^ In contrast, EVT is very effective in these patients, but is restricted to comprehensive stroke centers (CSC). aLVO patients who are first presented in a primary stroke center (PSC) therefore require subsequent transfer to a CSC, resulting in treatment delays and worse clinical outcomes.^6-8^

Treatment options for hemorrhagic stroke have been limited so far. However, it is becoming increasingly clear that fast initiation of treatment, including blood pressure management and reversal of coagulopathy, also improves outcomes in these patients.^9,10^ Although the exact role of surgery is still unclear, a recent meta-analysis showed that it is more effective when performed shorter after onset.^11^ Moreover, new techniques such as minimally invasive surgery are promising and also demonstrated better results when performed earlier.^11-15^ Similarly, timely neurosurgical interventions may be necessary in other types of intracranial hemorrhage (ICH) as well, including subarachnoid or traumatic hemorrhages.^9,10^

These neurosurgical interventions are also restricted to CSCs, which stresses the importance of prehospital recognition of ICH alongside aLVO stroke patients for direct transportation to a CSC. Previous studies have shown that certain demographic characteristics and clinical observations, e.g. advanced age, use of oral anticoagulation, decreased consciousness, vomiting and elevated blood pressure, are associated with ICH.^16-22^ However, sample sizes of these studies were generally small and they often assessed only a few features.

Furthermore, other studies demonstrated that it is difficult to specifically identify ICH patients based solely on clinical assessment, and that current triage scores are insufficient.^22-26^ Concerning the triage of aLVO patients, studies comparing several clinical scales demonstrated that the Rapid Arterial oCclusion Evaluation (RACE) score performs relatively well.^27-29^ However, it is yet unclear if the RACE score can also be used for recognition of ICH patients. Therefore, we aim to 1) assess the utility of the RACE score for prehospital identification of patients with ICH in addition to aLVO stroke, and 2) to compare the RACE to other demographic characteristics and clinical observations that have been shown to distinguish patients with ICH or aLVO stroke from patients with non-aLVO ischemic stroke, TIA or stroke mimic.

## METHODS

### Study population and data collection

We used data from the Leiden Prehospital Stroke Study (LPSS), a large prospective, multicenter, observational cohort study in two Emergency Medical Services (EMS) regions in the Netherlands. These regions encompass 4 PSCs and 3 CSCs, serving approximately 2 million inhabitants.^27^ The study included all patients ≥18 years old for whom an EMS-initiated acute stroke code was activated between July 2018 and October 2019. EMS paramedics were registered nurses with specialized training in ambulance care, who activated a stroke code based on a positive Face-Arm-Speech Time (FAST) test or other (focal) neurological symptoms at the discretion of the individual paramedics. Policy was to transport patients to the nearest hospital (PSC or CSC) if onset was <6 hours before presentation, and to the nearest CSC if onset was between 6 and 24 hours. Paramedics routinely documented patient characteristics and clinical observations in electronic transport records, including blood pressure, glucose level, assessment of consciousness with the AVPU (Alert/Verbal/Pain/Unresponsive) score and Glasgow Coma Scale, pupillary assessment, and presence of nausea/vomiting. For the LPSS an additional web-based application containing 10 to 13 structured clinical items derived from the National Institutes of Health Stroke Scale (NIHSS) was filled in for each patient.^30^ This enabled the prehospital reconstruction of several aLVO scales including the RACE. The RACE encompasses six clinical observations (facial palsy, arm and leg motor deficits, gaze deviation and either agnosia or aphasia), resulting in a score of 0 to 9 points. A RACE score of ≥5 points was considered positive.^29^ Corresponding electronic patient records from the hospitals were used to extract medical history, medication use, admission NIHSS score, data on neuroimaging including location of ICH and aLVO, final diagnosis after 3 months, and functional outcome after 3 months using the modified Rankin Scale (mRS).^31^ ICH were categorized based on the underlying cause and location as either primary hemorrhages, including deep, lobar or posterior fossa intraparenchymal hemorrhages, subarachnoid hemorrhages or intraventricular hemorrhages, or as secondary traumatic hemorrhages. aLVO was defined as an occlusion of the internal carotid artery, M1 or M2-part of the middle cerebral artery or A1 or A2-part of the anterior cerebral artery. Patients with missing transport records, unutilized web-based applications or missing hospital records were excluded. We used the STROBE guidelines for reporting this observational study (**Table S1**).^32^

### Statistical analysis

Patients were categorized into three groups based on their final diagnosis: 1) ICH; 2) aLVO stroke; or 3) other (non-aLVO stroke, TIA or stroke mimic). Independent t-tests and chi-square tests were used to compare baseline characteristics between the three groups. For our first aim, we calculated sensitivity, specificity, positive predictive value (PPV) and negative predictive value (NPV) of a positive RACE score for a diagnosis of ICH or aLVO stroke. To explore how use of the RACE for direct transportation to a CSC might influence patient allocation, we provided a hypothetical example in our cohort. For our second aim, we conducted multivariable logistic regression analyses to investigate associations of other demographic characteristics and prehospital EMS observations with a diagnosis of ICH or aLVO stroke, and to compare these with the RACE score. These characteristics and observations were selected based upon previous literature, and included age, sex, history of atrial fibrillation, use of oral anticoagulation, mean arterial blood pressure, glucose level, consciousness as measured with the AVPU and Glasgow Coma Scale, pupillary assessment, and presence of nausea/vomiting.^16-22,33^ We calculated adjusted odds ratios (aOR) with 95% confidence intervals (CI) to determine associations with a final diagnosis of ICH or aLVO stroke, with the RACE score as (a) a dichotomized variable (positive ≥5 points or negative <5 points) and (b) as a categorical variable ranging from 0 to 9 points. A 2-sided p-value of ≤0.05 was considered statistically significant.

### Missing data

For patients in whom one or more items of the RACE were untestable or missing, we assessed if the cut-off of ≥5 was already reached with the points scored in documented items (positive), or if the score would still be less than 5 even when assigning maximal scores to missing items (negative). Patients in whom it could not be determined if the RACE was positive or negative were excluded from analysis of sensitivity, specificity, PPV and NPV of the RACE. In case a history of atrial fibrillation or use of oral anticoagulation was not documented by EMS personnel, this was extracted from hospital records. For multivariable analyses, missing data including RACE scores were filled using multiple imputation by chained equations (MICE) with five imputations.^34^ Variables used during MICE constituted of all variables used in the multivariable analyses and the final diagnosis. We used Rubin’s rules to pool outcomes of the multivariable analyses from the five imputations.

### Outcomes

Primary outcomes were the sensitivity, specificity, PPV and NPV of the RACE score for ICH or aLVO stroke. Secondary outcome was the aOR of the RACE compared to other characteristics and observations in multivariable analysis.

## RESULTS

Of 2812 acute stroke codes activated, 808 (28.7%) were excluded because no web-based application was used by EMS (n=752), or hospital records (n=53) or transport records (n=3) were missing. In a previous analysis, patients in whom the application was not used had similar baseline characteristics, incidence of aLVO, and stroke severity, but slightly more often an ICH (12.0% vs. 7.4%) or stroke mimic (44.6% vs. 37.5%) compared with included patients.^27^ Of the 2004 included stroke code patients, 149 (7.4%) had an ICH, 153 (7.6%) an aLVO stroke, and 1702 (84.9%) other diagnoses (687 non-aLVO stroke, 262 TIA and 753 stroke mimic) (**Figure 1**). In total, 780 (38.9%) were first presented in a PSC and 1224 (61.1%) in a CSC. Of the 149 ICH patients, 130 (87.2%) had a primary hemorrhage of which 116 were intraparenchymal, 13 subarachnoid and 1 intraventricular, and 19 had a secondary traumatic hemorrhage. Thirty-eight (25.5%) were presented in a PSC and 111 (74.5%) in a CSC. Of the 153 aLVO stroke patients, 31 (20.3%) were first transported to a PSC, and 98 (64.1%) underwent EVT. Of these EVT-treated patients, 25 were initially presented in a PSC and subsequently transferred to a CSC (**Table 1**).

**Figure 1.**
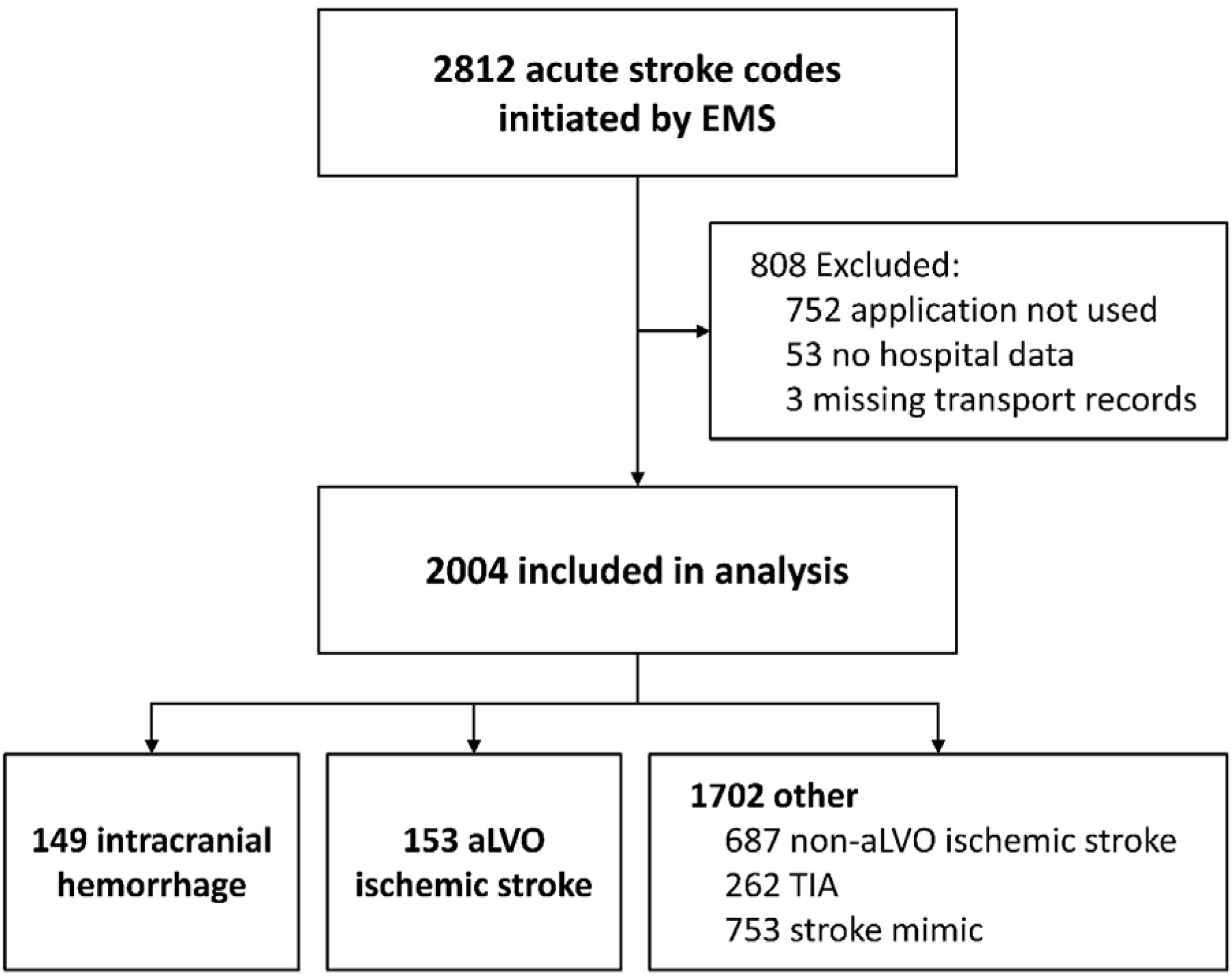
Flowchart of included patients. EMS = emergency medical services; aLVO = anterior circulation large vessel occlusion; TIA = transient ischemic attack

**Table 1.** Characteristics of patients with either intracranial hemorrhage, aLVO stroke or other diagnoses

|  | **Total (n=2004)** | **Intracranial hemorrhage (n=149)** | **aLVO stroke (n=153)** | **Other\* (n=1702)** |
| --- | --- | --- | --- | --- |
| **Demographic characteristics** |  |  |  |  |
| Age, mean (SD), years | 71.1 (14.9) | 73.7 (13.2) | 72.7 (13.0) | 70.7 (15.1) |
| Sex, male | 1020/2004 (50.9%) | 82/149 (55.0%) | 90/153 (58.8%) | 848/1702 (49.8%) |
| **Presentation** |  |  |  |  |
| PSC | 780/2004 (38.9%) | 38/149 (25.5%) | 31/153 (20.3%) | 711/1702 (41.8%) |
| CSC | 1224/2004 (61.1%) | 111/149 (74.5%) | 122/153 (79.7%) | 991/1702 (58.2%) |
| Pre-morbid mRS $\leq 2$ | 1566/1833 (85.4%) | 119/149 (79.9%) | 123/135 (91.1%) | 1324/1549 (85.5%) |
| Wake-up/unknown time of symptom onset | 326/1604 (20.3%) | 16/102 (15.7%) | 46/149 (30.9%) | 264/1353 (19.5%) |
| Onset-to-EMS-arrival time, median (IQR), minutes | 93 (32-276) | 91 (20-332) | 92 (17-305) | 93 (34-273) |
| Onset-to-door time, median (IQR), minutes | 119 (61-300) | 118 (51-352) | 115 (45-324) | 119 (64-300) |
| **Medical history & medication** |  |  |  |  |
| Atrial fibrillation | 272/1982 (13.7%) | 32/147 (21.8%) | 28/149 (18.8%) | 212/1686 (12.6%) |
| Intracranial hemorrhage | 83/1995 (4.2%) | 14/147 (9.5%) | 2/148 (1.3%) | 67/1698 (3.9%) |
| Ischemic stroke or TIA | 670/1984 (33.8%) | 36/146 (24.7%) | 32/150 (21.3%) | 602/1688 (35.7%) |
| Diabetes mellitus | 420/1983 (21.2%) | 23/147 (15.6%) | 36/150 (24.0%) | 361/1686 (21.4%) |
| Epilepsy | 106/2004 (5.3%) | 4/149 (2.7%) | 4/153 (2.6%) | 98/1702 (5.8%) |
| Use of oral anticoagulation | 336/1964 (17.1%) | 40/145 (27.6%) | 22/151 (14.6%) | 274/1668 (16.4%) |
| Use of antiplatelet medication | 689/1974 (34.9%) | 42/145 (29.0%) | 49/151 (32.5%) | 598/1678 (35.6%) |
| **EMS observations** |  |  |  |  |
| **Blood pressure, median (IQR), mm Hg** |  |  |  |  |
| Systolic | 160 (141-182) | 178 (155-197) | 155 (138-178) | 159 (141-181) |
| Diastolic | 90 (80-102) | 98 (86-110) | 90 (80-101) | 90 (80-101) |
| Glucose level, median (IQR), mmol/L | 6.5 (5.5-7.8) | 6.9 (5.7-8.4) | 6.6 (5.5-8.0) | 6.4 (5.5-7.8) |
| Abnormal AVPU score (Verbal/Pain/Unresponsive) | 283/2004 (14.1%) | 57/149 (38.3%) | 36/153 (23.5%) | 190/1702 (11.2%) |
| Abnormal Glasgow Coma Scale (<15 points) | 433/1995 (21.7%) | 65/148 (43.9%) | 49/153 (32.0%) | 319/1694 (18.8%) |
| Eyes – not open spontaneously (<4) | 169 (8.5%) | 42 (28.4%) | 21 (13.7%) | 106 (6.3%) |
| Motor – does not execute commands (<6) | 192 (9.6%) | 44 (29.7%) | 30 (19.6%) | 118 (7.0%) |
| Verbal – not oriented (<5) | 394 (19.7%) | 59 (39.9%) | 48 (31.4%) | 287 (16.9%) |
| Abnormal pupillary assessment (anisocoria or not reactive to light) | 46/2004 (2.3%) | 12/149 (8.1%) | 6/153 (3.9%) | 28/1702 (1.6%) |
| **Nausea/vomiting** | 370/700 (52.9%) | 39/60 (65.0%) | 14/34 (41.2%) | 317/606 (52.3%) |
| **RACE score positive ($\geq 5$ points)** | 195/1565 (12.5%) | 43/93 (46.2%) | 65/112 (58.0%) | 87/1360 (6.4%) |
| **In-hospital NIHSS score, median (IQR)** | 2 (0-5) | 11 (4-18) | 11 (5-17) | 1 (0-4) |
| **Imaging findings, treatment and outcome** |  |  |  |  |
| **Primary intracranial hemorrhage** |  | **130 (87.2%)** |  |  |
| Intraparenchymal |  | 116 (77.9%) |  |  |
| *Deep* |  | 65 (43.6%) |  |  |
| *Lobar* |  | 42 (28.2%) |  |  |
| *Posterior fossa (brainstem/cerebellum)* |  | 9 (6.0%) |  |  |
| Subarachnoid |  | 13 (8.7%) |  |  |
| Intraventricular |  | 1 (0.7%) |  |  |
| **Traumatic intracranial hemorrhage** |  | **19 (12.8%)** |  |  |
| Isolated subdural hematoma |  | 13 (8.7%) |  |  |
| Combined† |  | 6 (4.0%) |  |  |
| **Large vessel occlusion** | **173 (8.6%)** |  | **153** | **20** |
| Anterior circulation | 153 (7.6%) |  | 153 |  |
| *Internal carotid artery* |  |  | 16 (10.5%) |  |
| *Middle cerebral artery M1* |  |  | 75 (49.0%) |  |
| *Middle cerebral artery M2* |  |  | 58 (37.9%) |  |
| *Anterior cerebral artery* |  |  | 4 (2.6%) |  |
| Posterior circulation | 19 (0.9%) |  |  | 19 (95.0%) |
| Multiple occlusions‡ | 1 (0.1%) |  |  | 1 (0.5%) |
| **IVT** | 314 (15.7%) |  | 63/153 (41.2%) | 251/1702 (14.7%) |
| **EVT** | 98 (4.9%) |  | 98/153 (64.1%) |  |
| **transfer from PSC to CSC for EVT** |  |  | 25/98 (25.5%) |  |
| **mRS after 3 months $\leq 2$** | 470/787 (59.7%) | 27/103 (26.2%) | 45/112 (40.2%) | 398/572 (69.6%) |

\* Other diagnoses include non-aLVO stroke, TIA and stroke mimic.
† Combined: hemorrhage on multiple locations (e.g. epidural, subdural, subarachnoid and/or intraparenchymal).
‡ One patient had multiple occlusions in the anterior as well as posterior circulation.

Atrial fibrillation was more common in patients with ICH (21.8%) and aLVO stroke (18.8%) compared to other patients (12.6%, p<0.01 and p=0.03 respectively). Furthermore, ICH patients more often had a history of previous intracranial hemorrhage (respectively 9.5% vs. 1.3% and 3.9%, both p<0.01) and used oral anticoagulation (27.6% vs. 14.6% and 16.4%, both p<0.01) than patients with aLVO stroke or other diagnoses (**Table 1, Table S2**). Concerning EMS observations, ICH patients had higher blood pressure (median 178/98 vs. 155/90 and 159/90 mmHg respectively, both p<0.01), and more often an abnormal pupillary assessment (respectively 8.1% vs. 3.9% and 1.6%, p=0.13 and <0.01) and nausea/vomiting (65.0% vs. 41.2% and 52.3%, p=0.03 and 0.06) than patients with aLVO stroke or other diagnoses. Also, patients with ICH and to a lesser extent aLVO stroke more often had a decreased consciousness, scoring a Verbal/Pain/Unresponsive on the AVPU score (38.3% vs. 23.5% vs. 11.2%, p<0.01 for all differences) and less points on the Glasgow Coma Scale (<15 points: 43.9% and 32.0% vs. 18.8%; both p<0.01) than patients with other diagnoses (**Table 1, Table S2**). Statistical analysis of differences between the three groups can be found in **Table S2**, and further specification of characteristics of patients with non-aLVO stroke, TIA or stroke mimic in **Table S3**.

It was possible to determine a positive or negative RACE score in 1565 (78.1%) patients. Patients with ICH or aLVO stroke more often had a positive RACE score than patients with other diagnoses (46.2% and 58.0% vs. 6.4%, both p<0.01) (**Table 1, Table S2**). When combining the groups of ICH and aLVO stroke patients, a positive RACE score had a sensitivity of 52.7%, specificity of 93.6%, PPV of 55.4%, and NPV of 92.9% (**Table 2**). Hypothetically, if the RACE would have been applied to our cohort in an urban region with a relatively high proportion of CSCs and short distances between PSCs and CSCs, 9 of the 38 ICH patients and 17 of the 31 aLVO stroke patients (15 treated with EVT) who were first presented in a PSC would have benefited from direct allocation to a CSC. Thirty-nine patients with other diagnoses would have unnecessarily bypassed a closer PSC for transport to a CSC, including 18 non-aLVO stroke patients who received IVT.

**Table 2.** Sensitivity, specificity, PPV and NPV of a positive RACE score (≥5 points) for a diagnosis of intracranial hemorrhage or aLVO stroke

|  | **Sensitivity** | **Specificity** | **PPV** | **NPV** |
| --- | --- | --- | --- | --- |
| **ICH** vs. all other diagnoses (aLVO stroke, non-aLVO stroke, TIA and stroke mimic) | 46.2% | 89.7% | 22.1% | 96.4% |
| **ICH** vs. non-aLVO stroke, TIA and stroke mimic (without aLVO stroke) | 46.2% | 93.6% | 33.1% | 96.2% |
| **aLVO stroke** vs. all other diagnoses (ICH, non-aLVO stroke, TIA and stroke mimic) | 58.0% | 91.1% | 33.3% | 96.6% |
| **aLVO stroke** vs. non-aLVO stroke, TIA and stroke mimic (without ICH) | 58.0% | 93.6% | 42.8% | 96.4% |
| **ICH and aLVO stroke** vs. non-aLVO stroke, TIA and stroke mimic | 52.7% | 93.6% | 55.4% | 92.9% |

In multivariable analyses, male sex, higher mean arterial pressure and higher RACE scores were significantly associated with ICH or aLVO stroke (**Table 3, Figure S1**). A positive RACE score had strongest association with ICH or aLVO stroke (aOR 10.11, 95% CI 6.84-14.93) (**Table 3a**).

**Table 3.**
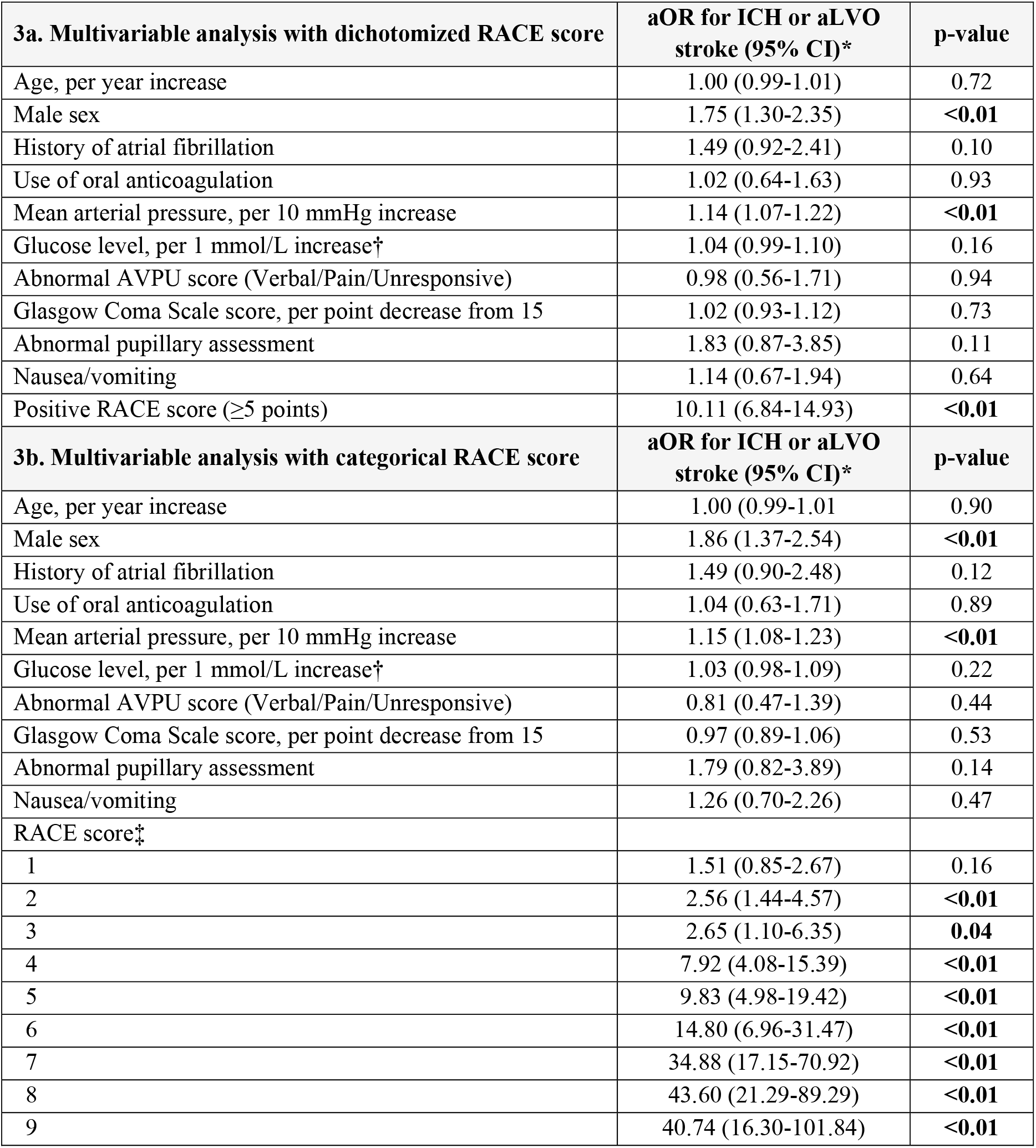
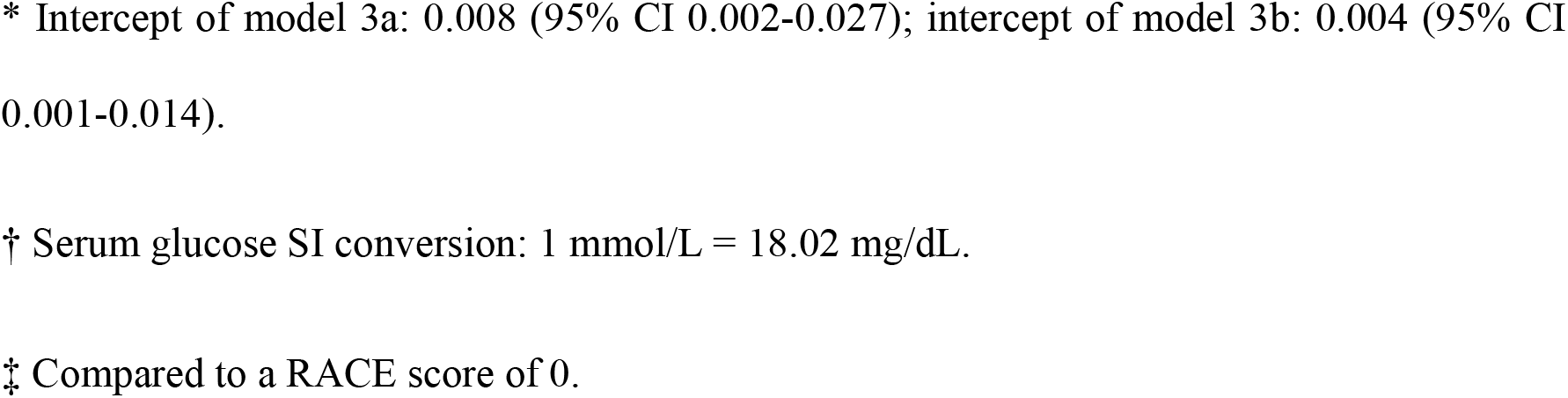
Multivariable regression analysis for diagnosis of ICH or aLVO stroke vs. other diagnoses

| **3a. Multivariable analysis with dichotomized RACE score** | **aOR for ICH or aLVO stroke (95% CI)\*** | **p-value** |
| --- | --- | --- |
| Age, per year increase | 1.00 (0.99-1.01) | 0.72 |
| Male sex | 1.75 (1.30-2.35) | **<0.01** |
| History of atrial fibrillation | 1.49 (0.92-2.41) | 0.10 |
| Use of oral anticoagulation | 1.02 (0.64-1.63) | 0.93 |
| Mean arterial pressure, per 10 mmHg increase | 1.14 (1.07-1.22) | **<0.01** |
| Glucose level, per 1 mmol/L increase† | 1.04 (0.99-1.10) | 0.16 |
| Abnormal AVPU score (Verbal/Pain/Unresponsive) | 0.98 (0.56-1.71) | 0.94 |
| Glasgow Coma Scale score, per point decrease from 15 | 1.02 (0.93-1.12) | 0.73 |
| Abnormal pupillary assessment | 1.83 (0.87-3.85) | 0.11 |
| Nausea/vomiting | 1.14 (0.67-1.94) | 0.64 |
| Positive RACE score ($\geq 5$ points) | 10.11 (6.84-14.93) | **<0.01** |
| **3b. Multivariable analysis with categorical RACE score** | **aOR for ICH or aLVO stroke (95% CI)\*** | **p-value** |
| Age, per year increase | 1.00 (0.99-1.01) | 0.90 |
| Male sex | 1.86 (1.37-2.54) | **<0.01** |
| History of atrial fibrillation | 1.49 (0.90-2.48) | 0.12 |
| Use of oral anticoagulation | 1.04 (0.63-1.71) | 0.89 |
| Mean arterial pressure, per 10 mmHg increase | 1.15 (1.08-1.23) | **<0.01** |
| Glucose level, per 1 mmol/L increase† | 1.03 (0.98-1.09) | 0.22 |
| Abnormal AVPU score (Verbal/Pain/Unresponsive) | 0.81 (0.47-1.39) | 0.44 |
| Glasgow Coma Scale score, per point decrease from 15 | 0.97 (0.89-1.06) | 0.53 |
| Abnormal pupillary assessment | 1.79 (0.82-3.89) | 0.14 |
| Nausea/vomiting | 1.26 (0.70-2.26) | 0.47 |
| RACE score‡ |  |  |
| 1 | 1.51 (0.85-2.67) | 0.16 |
| 2 | 2.56 (1.44-4.57) | **<0.01** |
| 3 | 2.65 (1.10-6.35) | **0.04** |
| 4 | 7.92 (4.08-15.39) | **<0.01** |
| 5 | 9.83 (4.98-19.42) | **<0.01** |
| 6 | 14.80 (6.96-31.47) | **<0.01** |
| 7 | 34.88 (17.15-70.92) | **<0.01** |
| 8 | 43.60 (21.29-89.29) | **<0.01** |
| 9 | 40.74 (16.30-101.84) | **<0.01** |

\* Intercept of model 3a: 0.008 (95% CI 0.002-0.027); intercept of model 3b: 0.004 (95% CI 0.001-0.014).
† Serum glucose SI conversion: 1 mmol/L = 18.02 mg/dL.
‡ Compared to a RACE score of 0.

## DISCUSSION

Our findings show that a positive RACE score is associated with both ICH and aLVO stroke, whilst the predictive value of other demographic characteristics and EMS observations is limited. The promising future treatment options for ICH and increasing use of EVT emphasize the growing need for adequate identification of stroke patients that require direct transportation to a CSC.^11-14,35,36^ This study demonstrates the potential of the RACE for optimizing prehospital triage and subsequently minimizing treatment delays and improving outcomes for both ICH and aLVO stroke patients.

Whilst outcomes of patients with ischemic stroke have significantly improved due to the impact of reperfusion therapies, treatment options for hemorrhagic stroke have remained limited and prognosis has not clearly changed over the last 20 years.^37^ However, results of recent studies investigating minimally invasive surgery in the acute phase are encouraging and multiple trials are ongoing (ENRICH, NCT02880878; DIST, NCT03608423; MIND, NCT03342664; INVEST, NCT02654015).^11-14^ This stresses the importance of prehospital identification of ICH patients alongside aLVO stroke patients. Previous studies that aimed to distinguish patients with ICH from other diagnoses based on clinical findings have been disappointing.^22-26^ However, our study suggests that the RACE score could help identify these patients. Although its sensitivity was lower for ICH than for aLVO stroke (46.2% vs. 58.0%), it is likely that this is caused by patients with smaller hemorrhages and concomitantly less deficits having a false negative RACE score. However, in these patients surgical interventions are less often indicated and transportation to a CSC less urgent. Furthermore, the limited prevalence of ICH and aLVO strokes in our population may well explain the relatively low PPV (55.4%).

Use of the RACE score in our cohort would have hypothetically implicated that 18 IVT-treated non-aLVO stroke patients bypassed a closer PSC, whilst 15 EVT-treated aLVO patients and 9 ICH patients would have been directly allocated to a CSC. Set aside the possible benefit for ICH patients, this may well have led to improved outcomes since the additional time required for an interhospital transfer is generally much higher than the additional transport time to a CSC, and a reduction in delay to EVT usually outweighs a similar time reduction to IVT.^7,8,38-40^

In multivariable analysis, most other variables were not significantly associated with ICH or aLVO stroke and may be of limited use for triage in clinical practice. This is in line with previous studies that reported poor results of scoring systems that tried to identify ICH patients based on demographic characteristics and clinical observations, which often also included atrial fibrillation, level of consciousness and nausea/vomiting.^22-26^ Surprisingly, a decreased consciousness (Verbal/Pain/Unresponsive on the AVPU score or decrease in points on the Glasgow Coma Scale) was not associated with ICH or aLVO stroke in multivariable analysis, whilst this was more common in these patients. However, we found that this was due to its strong correlation with the RACE score, which overshadowed and inversed the effect of a decreased consciousness.

Strengths of this study include the large sample size, number of characteristics assessed and prospective collection of data. Because of this, the overall amount of missing data was limited and extensive evaluation of characteristics was possible. Furthermore, the study design was very pragmatic, including all stroke code patients, using clinical observations from paramedics on-site, and analyzing the whole group of ICH and aLVO stroke patients, which represent all patients who require direct transportation to a CSC. This makes results well generalizable and useful for routine practice.

However, our study has some limitations. Firstly, some patients were excluded because the application was not used or hospital or transport records were missing. Those excluded had a slightly higher incidence of ICH, which may have underestimated the PPV and the hypothetical implications of use of the RACE score in our cohort. Secondly, it was not possible to reconstruct the RACE score in approximately 20% of patients due to one or more missing observations of its six clinical items. This may also have reduced the implications of the RACE in our hypothetical example, and we used MICE for completing data for multivariable analyses. Thirdly, a possible history of atrial fibrillation was not routinely documented in EMS transport records and therefore extracted from hospital records. In practice, it may be difficult to obtain this information in an emergency setting, although information concerning use of anticoagulation is generally well known by EMS. Fourthly, data on neuroimaging data, including the presence, location and size of ICH and occlusions, was also collected from hospital records and not reviewed by an independent core lab. Finally, although we focused on ICH and aLVO stroke patients, other patients may also benefit from direct transportation to a CSC, such as patients with a posterior circulation occlusion.^41^ However, this was beyond the scope of the current study.

## CONCLUSIONS

Our study shows that a positive RACE score is strongly associated with both ICH and aLVO stroke, while other demographic characteristics and prehospital observations have limited value for distinguishing these patients. This highlights the potential of the RACE for optimizing prehospital triage and subsequent allocation of stroke patients, and emphasizes the importance of its implementation in routine practice.

## Data Availability

In compliance with Dutch law, patient data cannot be made available, since participants were not informed during the opt-out procedure about the public sharing of their individual participant data in de-identified form. The syntax and output files of the statistical analyses can be made available from the corresponding author upon reasonable request.

## Non-standard Abbreviations and Acronyms

aLVO: anterior circulation large vessel occlusion
aOR: adjusted odds ratio
AVPU: Alert/Verbal/Pain/Unresponsive
CI: confidence interval
CSC: comprehensive stroke center
EMS: emergency medical services
EVT: endovascular thrombectomy
ICH: intracranial hemorrhage
IVT: intravenous thrombolysis
LPSS: Leiden Prehospital Stroke Study
mRS: modified Rankin Scale
NIHSS: National Institutes of Health Stroke Scale
NPV: negative predictive value
PPV: positive predictive value
PSC: primary stroke center
RACE: Rapid Arterial oCclusion Evaluation

## Acknowledgements

We thank all the participants, site personnel, and local staff for making this study possible.

## Author Contributions

IvdW and NK conceptualized the study. LD and DD acquired data. EvZ consulted in statistical analyses and LD, VG, JH and EvZ carried out analyses. LD had full access to all the data in the study and takes responsibility for its integrity and the data analysis. LD drafted the manuscript. All authors critically critically revised the manuscript and approved the final version.

## Ethical Approval and Informed Consent

The institutional review boards of the Leiden University Medical Center and of the participating hospitals approved the original LPSS study. The need for obtaining informed consent was waived because the extent of effort required by the large number of health care providers was disproportionate compared with the relatively limited sensitivity of the collected data and intrusion to the personal privacy. The original study was registered with ClinicalTrials.gov (identifier: NCT04442659).

## Supplemental Material

Tables S1–S3

Figure S1

